# Factors Influencing Timely Utilization of HIV Viral Load Test Results in HIV Care: A Qualitative Study at Kisenyi Health Centre IV, Kampala

**DOI:** 10.64898/2026.10.21.26366176

**Authors:** Miriam Nabukenya, Ivan Ssenyonga, John Bosco Alege

## Abstract

Timely utilization of HIV viral load (VL) test results is critical for effective monitoring of antiretroviral therapy (ART) and achieving viral suppression among people living with HIV. However, delays in utilizing VL results remain a challenge in many resource-limited settings. This study explored the factors influencing timely utilization of VL test results at Kisenyi Health Centre IV in Kampala, Uganda. A qualitative study design was employed using key informant interviews and structured facility observations. Eighteen healthcare workers involved in HIV care and viral load management participated in the study, including clinicians, laboratory personnel, data clerks, counsellors, and facility management staff. Data were collected using semi-structured interview guides and observation checklists. Interview transcripts were analysed using thematic analysis guided by the Performance of Routine Information System Management (PRISM) Framework. Three major themes emerged: technical, behavioral, and organizational factors. Technical barriers included unstable digital systems, limited computer access, unreliable internet connectivity, and fragmented health information platforms that delayed result retrieval. Behavioral factors included workload pressures, training gaps, and varying levels of staff motivation and accountability. Organizational challenges included staffing shortages, weak supervision, inconsistent feedback mechanisms, and occasional stock-outs of printing materials. Observational findings confirmed that manual printing and re-entry of VL results into electronic systems contributed to delays in clinical decision-making. Timely utilization of viral load results is influenced by a combination of technical, behavioral, and organizational factors. Strengthening digital system integration, improving health worker capacity, and enhancing facility-level management support could significantly improve VL result utilization and HIV treatment outcomes.

## Introduction

Human Immunodeficiency Virus (HIV) remains a major global public health challenge, particularly in sub-Saharan Africa, which continues to bear the highest burden of the epidemic [34]. Viral load (VL) testing is the gold standard for monitoring the effectiveness of antiretroviral therapy (ART) and plays a critical role in assessing treatment response, detecting treatment failure, and guiding clinical decision-making for people living with HIV. Routine viral load monitoring enables health providers to identify patients with unsuppressed viral loads and implement timely interventions such as adherence counselling, regimen optimization, or switching to second-line therapy. Evidence shows that sustained viral load suppression is strongly associated with improved immune recovery, reduced HIV transmission, and better long-term clinical outcomes among patients receiving ART.

Despite global progress in scaling up HIV treatment services, timely access to and utilization of viral load test results remain a challenge in many low- and middle-income countries. Delays in viral load testing, result reporting, and clinical utilization can negatively affect treatment monitoring and compromise patient outcomes. Studies conducted in several resource-limited settings have reported prolonged turnaround times for viral load results due to challenges in laboratory infrastructure, sample transportation systems, and health information management systems. In many settings, weak communication between laboratories and clinical units further contributes to delays in translating laboratory results into clinical action.

In response to these challenges, health systems increasingly rely on digital health information systems to improve laboratory result management and data utilization. Effective use of routine health information systems depends not only on the availability of technological infrastructure but also on the behavioral and organizational context within which health workers operate. The Performance of Routine Information System Management (PRISM) Framework provides a comprehensive approach for understanding how technical, behavioral, and organizational factors interact to influence the performance and utilization of routine health information systems. According to this framework, the effectiveness of health information systems is shaped by factors such as system design, user competence, staff motivation, organizational support, and data-use culture.

In sub-Saharan Africa, several studies have highlighted barriers to the effective utilization of viral load results, including limited digital infrastructure, inadequate laboratory capacity, shortages of trained personnel, and weak integration between laboratory and clinical information systems. In Uganda, the scale-up of routine viral load testing has significantly improved monitoring of ART programs. However, challenges persist in ensuring that viral load results are accessed and utilized in a timely manner to inform clinical management. Delays in accessing results may limit the ability of healthcare providers to promptly identify patients with treatment failure and implement appropriate interventions.

Although previous studies in Uganda have examined viral load testing coverage and laboratory turnaround times, limited evidence exists on the broader health system factors that influence the timely utilization of viral load results at the facility level. Understanding these factors is essential for strengthening routine health information systems and improving HIV treatment outcomes. Exploring the experiences and perspectives of healthcare workers involved in viral load management can provide valuable insights into operational barriers and opportunities for improving the use of laboratory data in patient care.

Therefore, this study aimed to explore the technical, behavioral, and organizational factors influencing the timely utilization of HIV viral load test results at Kisenyi Health Centre IV in Kampala, Uganda. The findings are expected to inform strategies for improving viral load result management, strengthening routine health information systems, and enhancing the effectiveness of HIV treatment monitoring programs

## Materials and Methods

### Study Design

This was a health facility-based, cross-sectional qualitative study conducted at Kisenyi Health Centre IV (HCIV) in Kampala, Uganda. The study aimed to explore factors influencing the timely utilization of HIV viral load (VL) test results to optimize HIV treatment outcomes. A qualitative approach using inductive and deductive thematic analysis was employed. The study was guided by the PRISM (Performance of Routine Information System Management) analytical framework of Data Demand and Use (Measure Evaluation, USAID, 2020), focusing on behavioral, technical, and organizational factors affecting data use.

### Study Setting

Kisenyi HCIV is one of eight government health facilities managed by the Kampala Capital City Authority. It is a high-volume facility with a total of 7,405 clients actively on antiretroviral therapy (ART) by June 2024. The facility provides services through several clinics, including Antenatal Care, Outpatient Department (OPD), Dental Clinic, and ART Clinic, where all HIV-positive clients are managed.

### Study Population

The study population comprised healthcare workers directly or indirectly involved in ART service delivery. Key participants included facility-in-charge officers, ICT/data/M&E officers, clinical officers, nurses, pharmacists, and laboratory technologists involved in HIV care.

### Sampling Procedure

Non-probability consecutive sampling was used to recruit participants. Eligible participants were invited consecutively based on their availability and role until data saturation was achieved. Two research assistants, together with the principal investigator, conducted in-depth interviews and observations of clinic workflows. Saturation was determined when no new information emerged from interviews and observations.

### Sample Size

Sample size was guided by informational redundancy (Glaser & Strauss, 2017). Approximately 20 participants were selected for key informant interviews, and 10 observation sessions were conducted. Participants were purposively selected based on their ability to provide rich, relevant information regarding the research objectives.

### Data Collection Methods

#### Key Informant Interviews (KIIs)

In-depth, one-on-one interviews were conducted using a structured guide based on literature review. The guide had two sections: (1) participant background and (2) open- and closed-ended questions exploring knowledge, practices, and perceptions regarding the utilization of HIV VL results.

Data was collected from 3^rd^ October 2025 and concluded on 8^th^ October 2025

#### Observations

A structured observation checklist was used to systematically document clinic workflows, including patient triage, VL sample collection, result management, and utilization. Observational data were triangulated with interview data to enhance credibility.

### Data Management and Analysis

Interviews were audio-recorded, transcribed verbatim, and checked for accuracy. Data were analyzed using thematic framework analysis, combining inductive coding for emergent themes and deductive coding aligned with the PRISM framework. Themes were categorized under behavioral, technical, and organizational factors. Thematic analysis followed a systematic process of coding, categorizing, and interpreting patterns to derive insights relevant to timely utilization of HIV VL results (Luborsky & Rubinstein, 1995).

### Ethical Considerations

Ethical approval was obtained from the Research Ethics Committee of Clarke International University rec number CLARKE-2025-1594, and administrative approval from Kampala Capital City Authority. Written informed consent was obtained from all participants. Participation was voluntary, and confidentiality of responses was maintained. Data were stored securely and used solely for the purpose of this study.

### Dissemination of Findings

The study findings were disseminated through a final report submitted to Clarke International University Library, management of Kisenyi HCIV, and the Ministry of Health. Additionally, a manuscript will be prepared for publication in a peer-reviewed journal in the field of HIV/AIDS or infectious diseases.

## Results

### Participant Characteristics

A total of 18 key informants participated in the study, representing different professional cadres involved in HIV care and viral load (VL) management at Kisenyi Health Centre IV. Participants included clinicians, laboratory personnel, data clerks, counsellors, and a facility manager. The sample consisted of 10 females and 8 males, indicating relatively balanced gender representation.

Clinicians (medical officers, clinical officers, and nurses) constituted the largest group of participants (39%), followed by laboratory personnel (22%), data clerks and counsellors (17% each), and one facility manager (5%). Most participants had substantial experience in HIV care, with 61% reporting 6–12 years of work experience, while 39% had between 2–5 years of experience.

Although several participants had received training on viral load management and digital systems such as the Uganda Electronic Medical Records (UGEMR) system, others relied mainly on on-the-job learning. Differences in training exposure, professional roles, and years of experience influenced how staff interacted with viral load results and responded to delays in result utilization.

### Thematic Findings

Analysis of interview transcripts and observational data produced three major themes influencing the timely utilization of viral load test results:

Technical factors
Behavioral factors
rganizational factors

These themes align with the Performance of Routine Information System Management (PRISM) Framework, which emphasizes the interaction between technical systems, human behavior, and organizational context in the use of health information.

### Technical Fatcors of Viral Load Result Utilization

Technical factors emerged as the most frequently cited barriers affecting timely access to viral load results. Participants highlighted challenges related to limited access to digital infrastructure, unstable systems, and delays in result flow.

### Access to Technology and Digital Tools

Access to computers and digital platforms influenced how quickly staff could retrieve viral load results. Participants reported that limited computer availability and restricted system access often delayed result retrieval.

One participant explained:

“Only one computer in the laboratory is used to access results, so if it is busy or down, we have to wait until it is fixed before results can be printed.” (Data unit staff)

### System Functionality and Reliability

Participants described frequent system interruptions, including power outages and unreliable internet connectivity, which prevented timely printing and recording of viral load results.

“When the VL dashboard fails to load or power goes off, we cannot print results for patients.” (Laboratory staff)

### Sample Transportation and Result Flow

Although routine sample transportation was generally functional, participants reported long turnaround times from central testing laboratories, which delayed clinical decision-making.

“Sometimes we keep re-bleeding patients because results take months to come back.” (Clinician)

### Technical Support and Infrastructure

Limited technical support and fragmented digital systems further complicated access to results. Participants recommended integrating laboratory systems and improving technical maintenance.

### Behavioral Factors

Behavioral factors related to staff attitudes, motivation, accountability, and patient behavior also influenced the timely use of viral load results.

### Accountability and Responsibility

Participants reported that viral load result follow-up was largely dependent on a single VL focal person. When this individual was absent, results often remained unreviewed.

“Sometimes results come but no one follows up immediately when the VL focal person is away.” (Counsellor)

### Staff Attitudes and Workload

Heavy workload and staffing shortages reduced the ability of health workers to actively track viral load results.

“We were three people in this room before, but now I handle all the work alone.” (Counsellor)

### Training and Competence

Several participants reported limited training on viral load result interpretation, particularly for low-level viraemia.

“Not everyone knows how to interpret high or low viral load results.” (Clinician)

### Patient Behavior

Patient-related factors also influenced follow-up actions. Some patients delayed returning for counselling or denied unsuppressed viral load results.

“Some patients believe prayer will cure them, so they delay returning for follow-up.” (Counsellor)

### Organizational Factors

Organizational challenges were also identified as barriers to timely viral load result utilization.

### Availability of Resources

Participants reported frequent shortages of printing materials, which prevented immediate access to results even when they were available electronically.

“Sometimes results are ready online, but we cannot print them because there is no paper.” (Laboratory staff)

### Leadership and Management Support

Although management occasionally reviewed viral load performance, regular monitoring meetings were not consistently held.

### Staffing and Workload

Understaffing forced staff members to perform multiple roles, reducing the time available for reviewing viral load results.

### Standard Operating Procedures

Participants also reported limited awareness of standard operating procedures guiding viral load result management.

### Observational Findings

Facility observations confirmed many of the issues raised during interviews.

Health workers generally demonstrated strong commitment to reviewing viral load results during patient consultations. However, several operational barriers were observed, including intermittent system access, inconsistent documentation practices, and reliance on manual printing and re-entry of results into the electronic medical record system.

Observations also revealed that multiple health information systems were used simultaneously, including the Uganda EMR, Viral Load Dashboard, and Africa Laboratory Information System, which created inefficiencies in data flow.

## Discussion

### Summary of Key Findings

This study identified technical, behavioral, and organizational factors affecting the timely utilization of viral load test results at Kisenyi Health Centre IV. Technical barriers, including system instability, limited digital infrastructure, and fragmented information systems, were the most frequently reported challenges. Behavioral factors such as workload pressure, training gaps, and staff motivation also influenced result utilization, while organizational factors including staffing shortages and weak supervision further constrained timely clinical action.

These findings align with global research demonstrating that weak laboratory information systems and poor interoperability between digital health platforms can significantly delay clinical access to laboratory data. Studies in low- and middle-income countries have shown that fragmented digital systems often lead to delays in translating laboratory results into treatment decisions.

Additionally, global evidence highlights the importance of health worker motivation and organizational support in improving the use of routine health data. Even when health workers possess adequate knowledge, systemic constraints such as heavy workload and limited institutional support may prevent timely use of clinical information.

The findings are also consistent with research conducted across sub-Saharan Africa, where prolonged viral load turnaround times remain a common challenge. Studies in Uganda and Ethiopia have documented delays related to weak laboratory networks, equipment shortages, and limited digital infrastructure.

Research across African health systems also shows that understaffing and heavy workload reduce the ability of healthcare workers to actively track and utilize laboratory results. Similar to the present study, facilities with stronger leadership and routine performance monitoring tend to demonstrate faster clinical responses to viral load results.

### Policy and Program Implications

Improving the timely utilization of viral load results requires strengthening multiple components of the health system. Integrating laboratory information systems with electronic medical records would reduce the need for manual result printing and re-entry. In addition, continuous training and mentorship programs are necessary to improve health worker capacity in interpreting viral load results and using digital health systems.

Facility leadership should also strengthen performance monitoring and supervision mechanisms to ensure accountability in viral load result follow-up. Improved coordination between laboratories and ART clinics could further enhance communication and reduce delays in clinical decision-making.

### Future Research

Future studies should evaluate interventions aimed at improving digital system integration and reducing viral load turnaround time. Multi-facility studies across Uganda would also help determine whether the challenges observed at Kisenyi Health Centre IV reflect broader systemic issues within the national HIV program.

**Figure 1:**
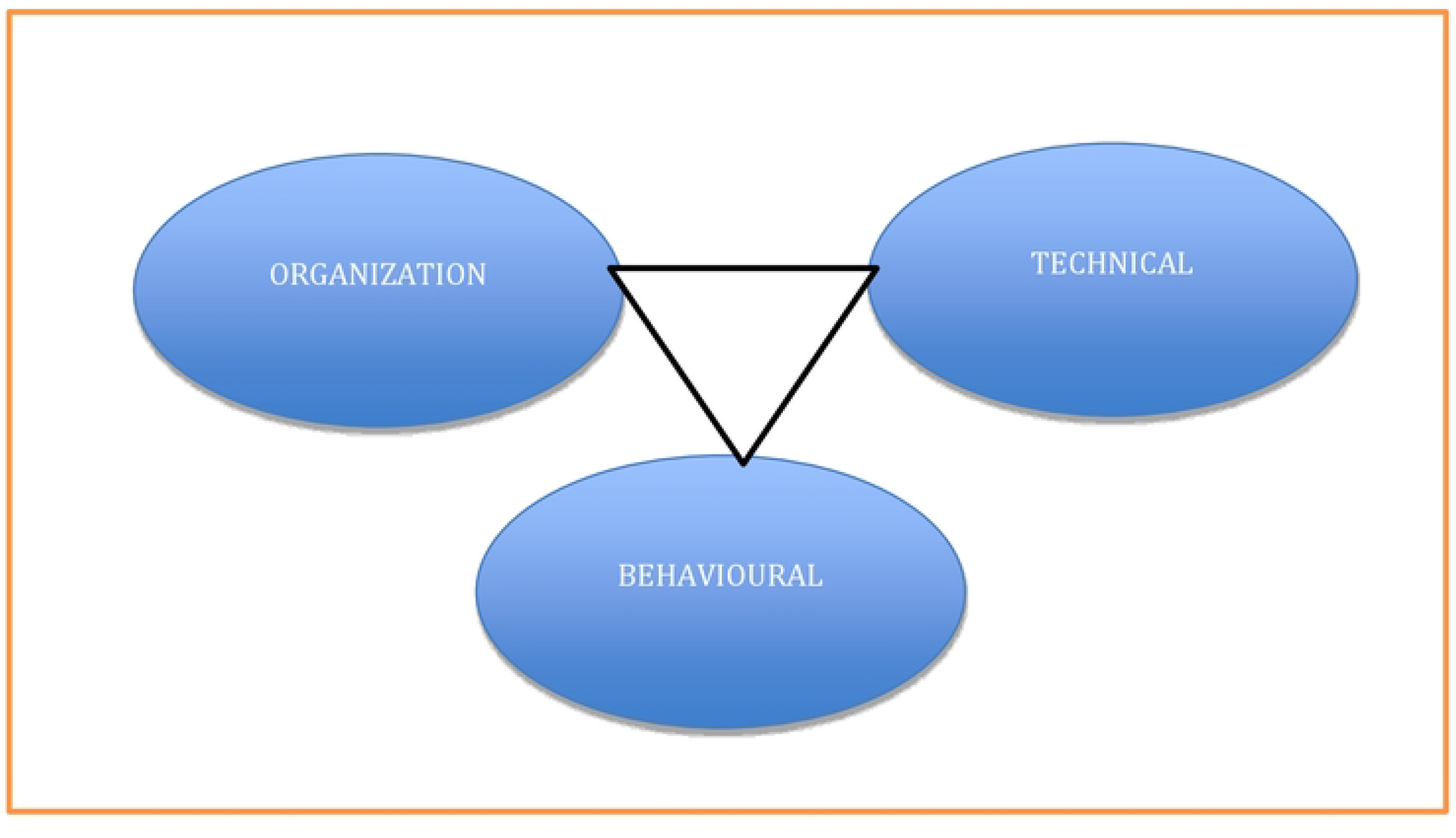
PRISM Analytical Framework of Data Demand and Use. Description: This figure illustrates the PRISM framework highlighting the interaction between organizational, technical, and behavioral determinants influencing data demand and use in health systems.

**Figure 2:**
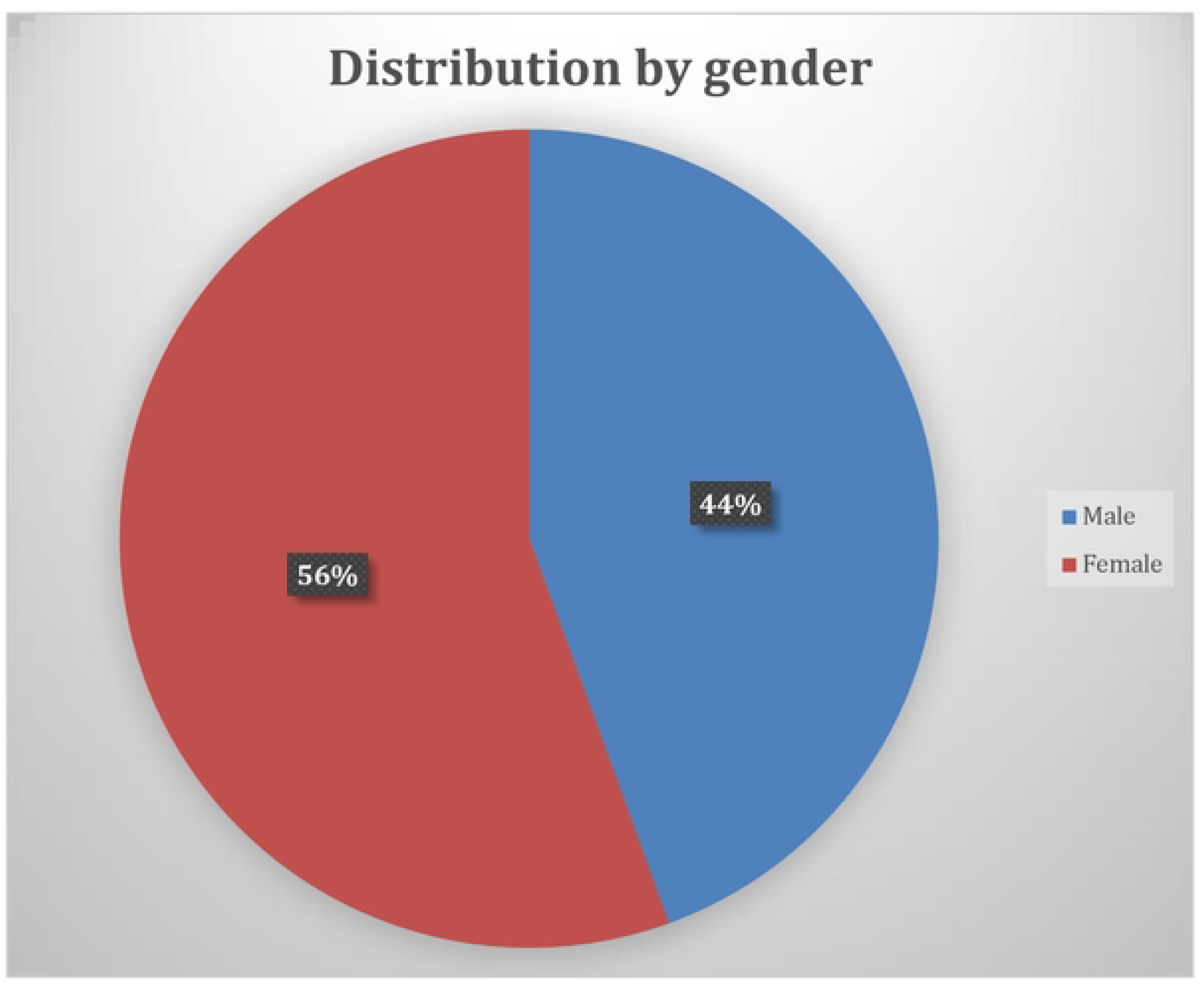
Distribution by Gender of Participants. Description: This figure presents the gender distribution of study participants, showing a relatively balanced representation between male and female respondents.

**Figure 3:**
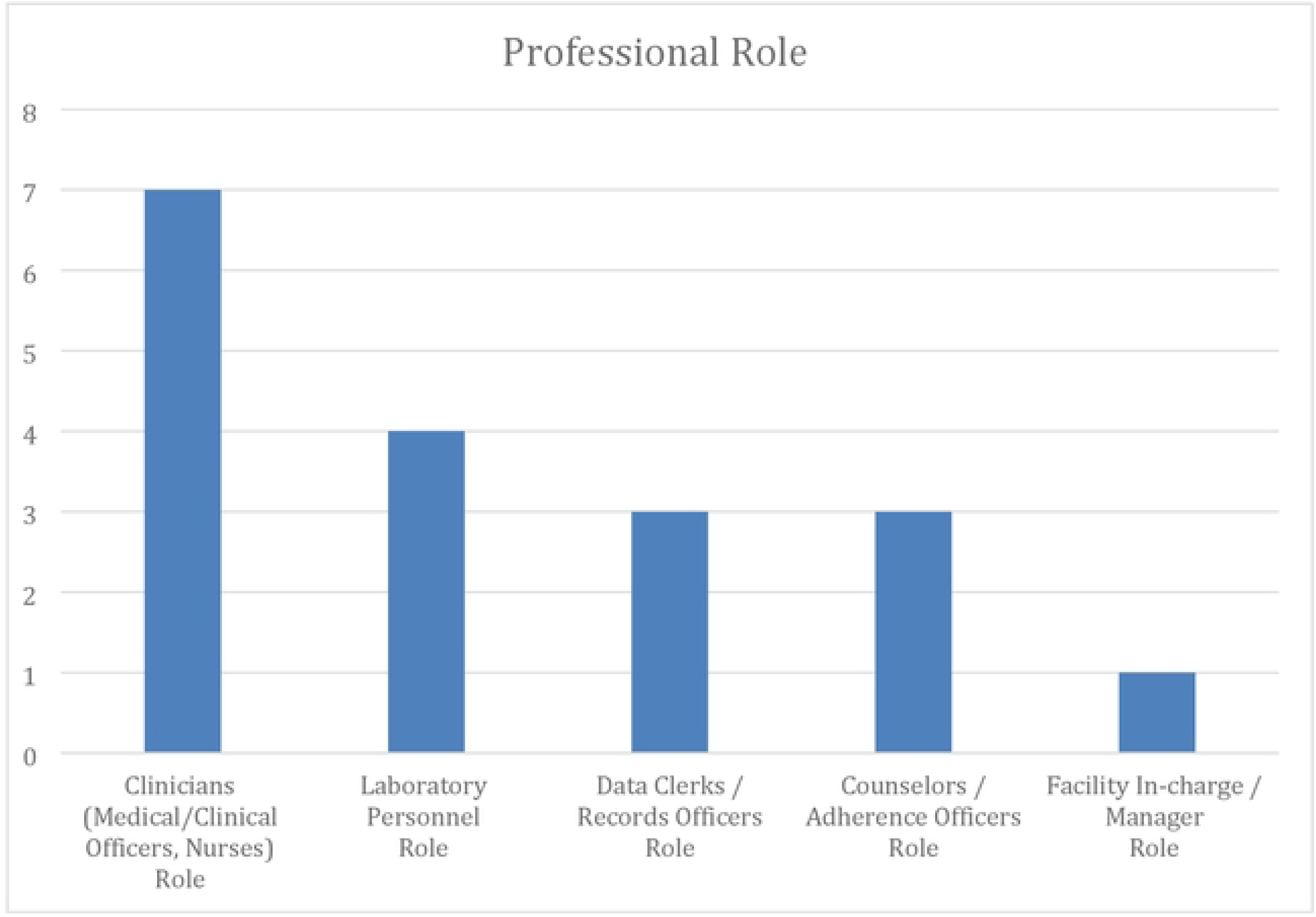
Professional Role of Participants. Description: This figure shows the distribution of participants by professional role, including clinicians, laboratory personnel, data clerks, counselors, and management

## Data Availability Statement

The datasets generated and analyzed during the current study are not publicly available due to confidentiality agreements with study participants but are available from the corresponding author upon reasonable request.

## Ethics Statement

Ethical approval for this study was obtained from the clerk international University Research Ethics Committee. Administrative permission to conduct the study was granted by Kisenyi Health Centre IV and the Kampala Capital City Authority (KCCA). Written informed consent was obtained from all participants prior to data collection. Participants were assured of confidentiality and anonymity throughout the study.

## Author Contributions

Conceptualization: Miriam Nabukenya

Methodology: Miriam Nabukenya

Data Collection: Miriam Nabukenya

Formal Analysis: Miriam Nabukenya

Writing Original Draft: Miriam Nabukenya

Writing Review & Editing: Ivan Ssenyonga

Supervision: John Bosco Alege

## Acknowledgments

I thank the management of Kisenyi HCIV and the entire ART clinic currently referred to as chronic care department for participating in my study clerk international University

